# Prior diagnoses and medications as risk factors for COVID-19 in a Los Angeles Health System

**DOI:** 10.1101/2020.07.03.20145581

**Authors:** Timothy S Chang, Yi Ding, Malika K Freund, Ruth Johnson, Tommer Schwarz, Julie M Yabu, Chad Hazlett, Jeffrey N Chiang, Ami Wulf, UCLA Health Data Mart Working Group, Daniel H Geschwind, Manish J Butte, Bogdan Pasaniuc

## Abstract

With the continuing coronavirus disease 2019 (COVID-19) pandemic coupled with phased reopening, it is critical to identify risk factors associated with susceptibility and severity of disease in a diverse population to help shape government policies, guide clinical decision making, and prioritize future COVID-19 research. In this retrospective case-control study, we used de-identified electronic health records (EHR) from the University of California Los Angeles (UCLA) Health System between March 9^th^, 2020 and June 14^th^, 2020 to identify risk factors for COVID-19 susceptibility (severe acute respiratory distress syndrome coronavirus 2 (SARS-CoV-2) PCR test positive), inpatient admission, and severe outcomes (treatment in an intensive care unit or intubation). Of the 26,602 individuals tested by PCR for SARS-CoV-2, 992 were COVID-19 positive (3.7% of Tested), 220 were admitted in the hospital (22% of COVID-19 positive), and 77 had a severe outcome (35% of Inpatient). Consistent with previous studies, males and individuals older than 65 years old had increased risk of inpatient admission. Notably, individuals self-identifying as Hispanic or Latino constituted an increasing percentage of COVID-19 patients as disease severity escalated, comprising 24% of those testing positive, but 40% of those with a severe outcome, a disparity that remained after correcting for medical co-morbidities. Cardiovascular disease, hypertension, and renal disease were premorbid risk factors present before SARS-CoV-2 PCR testing associated with COVID-19 susceptibility. Less well-established risk factors for COVID-19 susceptibility included pre-existing dementia (odds ratio (OR) 5.2 [3.2-8.3], p=2.6 × 10^−10^), mental health conditions (depression OR 2.1 [1.6-2.8], p=1.1 × 10^−6^) and vitamin D deficiency (OR 1.8 [1.4-2.2], p=5.7 × 10^−6^). Renal diseases including end-stage renal disease and anemia due to chronic renal disease were the predominant premorbid risk factors for COVID-19 inpatient admission. Other less established risk factors for COVID-19 inpatient admission included previous renal transplant (OR 9.7 [2.8-39], p=3.2×10^−4^) and disorders of the immune system (OR 6.0 [2.3, 16], p=2.7×10^−4^). Prior use of oral steroid medications was associated with decreased COVID-19 positive testing risk (OR 0.61 [0.45, 0.81], p=4.3×10^−4^), but increased inpatient admission risk (OR 4.5 [2.3, 8.9], p=1.8×10^−5^). We did not observe that prior use of angiotensin converting enzyme inhibitors or angiotensin receptor blockers increased the risk of testing positive for SARS-CoV-2, being admitted to the hospital, or having a severe outcome. This study involving direct EHR extraction identified known and less well-established demographics, and prior diagnoses and medications as risk factors for COVID-19 susceptibility and inpatient admission. Knowledge of these risk factors including marked ethnic disparities observed in disease severity should guide government policies, identify at-risk populations, inform clinical decision making, and prioritize future COVID-19 research.

## Introduction

Global cases of novel coronavirus disease 2019 (COVID-19) reached 9.8 million in June 2020, with the United States accounting for 2.4 million cases^1,2^. While still in the midst of the first acute phase of the pandemic^2–4^, knowledge of risk factors associated with COVID-19 susceptibility and severity can shape government policies, identify at-risk populations, guide clinical decision making, and prioritize future COVID-19 research. Existing studies have identified risk factors in hospitalized COVID-19 patients that are associated with death, intensive care unit admission or ventilation. Hypertension, cardiovascular disease, renal disease, and diabetes have consistently been identified as risk factors for a severe outcome in hospitalized patients in China, Italy and the United States^5–14^. On the other hand, there have been limited analyses on risk factors that confer an increased susceptibility for being diagnosed with COVID-19 (severe acute respiratory syndrome coronavirus 2 (SARS-CoV-2) positivity), and inpatient admission, both of which are important stages of the COVID-19 disease spectrum. In addition, data collection in previous studies were predominantly performed via standardized forms from electronic health records (EHR)^8,14,15^ or registry data reported to centralized coordinating centers^7,12^. These data are standardized, less detailed, and have limited access to records from previous encounters compared to direct EHR extraction^6,16^.

In addition, certain COVID-19 risk factors may be emphasized based on study design and vary due to the geographical and population context^17^. Within California, Los Angeles County has seven times more COVID-19 cases (93,000) compared to the second ranked county^18^. Los Angeles comprises a racially and ethnic diverse population with 48% self-identified Hispanic or Latinos, 15% Asians, and 9% Black or African Americans spread over 4,000 square miles^19^. With hospitals and clinics predominantly located in Los Angeles County, the University of California, Los Angeles (UCLA) Health System cares for individuals from this community. In this retrospective study, our objective was to identify pre-existing risk factors for COVID-19 diagnosis susceptibility, inpatient admission, and severe outcome through the automated EHR extraction of a large metropolitan medical system^20^. We identified several less established risk factors and also confirmed many known demographic and clinical COVID-19 risk factors.

## Results

### Study Subjects

The UCLA Health System includes two hospitals (520 and 281 inpatient beds) and 210 primary and specialty outpatient locations predominantly located in Los Angeles County. We leveraged an extract of the de-identified electronic health records from the UCLA Health System known as the Data Discovery Repository (DDR), which contains longitudinal records for more than 1.5 million patients since 2013.

This retrospective analysis included individuals who were tested for SARS-CoV-2 via RT-PCR within the UCLA Health System between March 9, 2020 and June 14, 2020, where the first positive SARS-CoV-2 PCR test was on March 12, 2020. Of the 26,602 individuals PCR tested for SARS-CoV-2 (Tested) (55% female, 44% male; mean (standard deviation, SD) 49 (20) years old), 992 were COVID-19 positive (3.7% of Tested; 51% female, 48% male; mean (SD) 49 (20) years old), 220 were admitted in the hospital (Inpatient) (22% of COVID-19 positive; 43% female, 56% male; mean (SD) 62 (21) years old) and 77 were treated in the intensive care unit or required intubation (Severe) (7.7% of COVID-19 positive, 35% of Inpatient; 38% female, 61% male; mean (SD) 56 (20) years old) (Figure 1).

**Figure 1.**
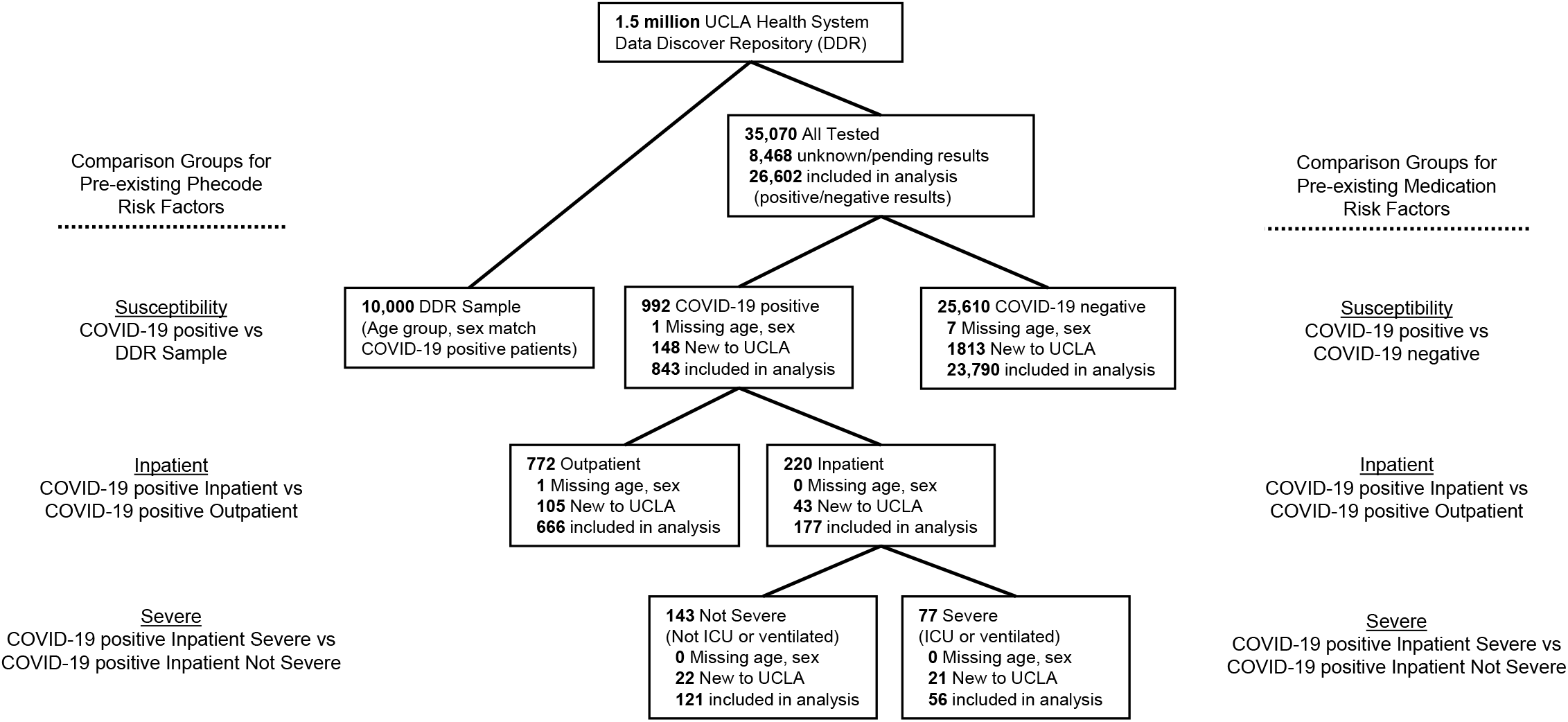
Flow diagram of subjects included in COVID-19 susceptibility, inpatient and severe analyses for pre-existing phecodes and medications

### Demographics

Compared to all tested patients, COVID-19 positive patients were significantly more likely to be male (48% male COVID-19 positive vs. 44% male Tested; odds ratio (OR) = 1.2 [1.0, 1.3]; p=0.02), and adults (2% <18 years old COVID-19 positive vs. 6% <18 years old Tested; OR =0.40 [0.26, 0.57]; p<0.001). Compared to COVID-19 positive patients, inpatients were more likely male (56% male Inpatients vs. 48% male COVID-19 positive; OR = 1.5 [1.1, 2.0]; p=6.0 × 10^−3^) and older than 65 years old (49% >65 years old Inpatient vs. 20% >65 years old COVID-19 positive; OR = 6.9 [4.9, 9.7]; p<0.001) (Table 1). These findings confirm previous reports that males and non-young individuals face higher risks of severe disease^21–23^.

**Table 1:** Demographics of COVID-19 patient groups. (↓ /↑) indicates a statistically significant negative/positive association (p<0.05) of the demographic and two patient groups. For age group and sex, percentages of Severe was compared to Inpatient, Inpatient was compared to COVID-19 positive, and COVID-19 positive was compared to Tested. Significant association of race and ethnicity for Severe compared Inpatient Severe vs. Inpatient Not Severe; for Inpatient compared COVID-19 positive Inpatient vs COVID-19 positive Outpatient; and for COVID-19 positive compared COVID-19 positive vs. COVID-19 negative while controlling for age group and sex.

|  | Demographic | Severe (77) | Inpatient (220) | COVID-19 positive (992) | Tested (26,602) |
| --- | --- | --- | --- | --- | --- |
| Age Group | < 18 years | 4 (5%) | 8 (3%) | 26 (2%)↓ | 1689 (6%) |
|  | 19-25 years | 2 (2%) | 5 (2%)↓ | 61 (6%) | 1458 (5%) |
|  | 26-55 years | 29 (37%) | 61 (27%)↓ | 547 (55%)↑ | 12,589 (47%) |
|  | 56-65 years | 15 (19%) | 37 (16%) | 152 (15%) | 4507 (16%) |
|  | > 65 years | 27 (35%)↓ | 109 (49%)↑ | 205 (20%)↓ | 6351 (23%) |
| Sex | Female | 30 (38%) | 96 (43%)↓ | 512 (51%)↓ | 14,679 (55%) |
|  | Male | 47 (61%) | 124 (56%)↑ | 479 (48%)↑ | 11,915 (44%) |
| Race | White or Caucasian | 26 (33%)↓ | 106 (48%)↓ | 428 (52%)↓ | 14,620 (54%) |
|  | Black or African American | 12 (15%) | 23 (10%) | 81 (8%)↑ | 1865 (7%) |
|  | Asian | 9 (11%) | 22 (9%) | 78 (7%) | 2495 (9%) |
|  | American Indian or Alaska Native | 0 (0%) | 0 (0%) | 4 (0%) | 89 (0%) |
|  | Native Hawaiian or Other Pacific Islander | 0 (0%) | 0 (0%) | 0 (0%) | 55 (0%) |
|  | Other Race | 29 (37%) | 68 (30%)↑ | 227 (22%) | 4551 (17%) |
|  | Unknown Race | 1 (1%) | 1 (0%) | 173 (17%) | 2919 (10%) |
| Ethnicity | Hispanic or Latino | 31 (40%) | 73 (33%)↑ | 245 (24%)↑ | 4338 (16%) |
|  | Not Hispanic or Latino | 44 (57%) | 143 (64%)↓ | 584 (58%)↓ | 19,254 (72%) |
|  | Unknown Ethnicity | 2 (2%) | 4 (2%) | 162 (16%) | 3002 (11%) |

However, in our dataset, both older (>65 years old) and intermediate (26-55 year old) age groups had an increased risk of a severe outcome depending on the comparison group. The percentage of Severe individuals >65 years old (35%) was higher than the percentage of COVID-positive individuals >65 years old (20%), but lower than the percentage of Inpatient individuals >65 years old (35% >65 years old Severe vs. 49% >65 years old Inpatients; OR = 0.41 [0.23, 0.71]; p=1.6 × 10^−3^). In contrast, the percentage of Severe individuals 26-55 years old (37%) was lower than the percentage of COVID-positive individuals 26-55 years old (55%), but higher than the percentage of Inpatient individuals 26-55 years old (37% 26-55 year old Severe vs. 27% 26-55 year old Inpatient; OR = 2.1 [1.1, 3.8]; p=0.016) (Table 1). These data show that the intermediate age group has a lower risk of inpatient admission, but higher risk of a severe outcome if admitted.

Self-identified Hispanic or Latino individuals constituted an increasing percentage of COVID-19 patients as disease severity increased. While Hispanic or Latino individuals comprised 16% of the Tested population, they comprised 24% of COVID-19 positive individuals (24% COVID-19 positive vs. 16% Tested; OR = 2.0 [1.7, 2.3]; p<0.001), 33% of Inpatient individuals (33% Inpatient vs. 24% COVID-19 positive; OR = 2.1 [1.4, 3.1]; p<0.001), and 40% of Severe individuals (40% Severe vs. 33% COVID-19 positive; OR = 1.3 [0.7, 2.4]; p=0.43) (Table 1). These findings show that self-identified Hispanic or Latino individuals had a higher risk of testing positive, and that when positive, suffered from more severe COVID-19 disease.

To evaluate if co-morbidities contributed to the disproportionate COVID-19 disease severity in Hispanic or Latino individuals, we next corrected for patient histories of known COVID-19 risk factors such as type 2 diabetes, hypercholesterolemia, hypertension, and chronic renal disease (Methods, Table S1), many of which are known to disproportionately affect Hispanic or Latino individuals^24,25^. The odds ratio for having a positive test remained >2.0 in Hispanic or Latino individuals, and for having a Severe disease course remained >2.7 (Table S2). These findings support that individuals who self-identify as Hispanic or Latino have risk factors, other than medical, that increase their risk of having COVID-19 disease and a severe outcome^26^.

Overall, individuals self-identifying as non-white (Asian, Black or African American, American Indian or Alaska Native, Native Hawaiian or Other Pacific Islander, Other Race) showed increased COVID-19 susceptibility (39% COVID-19 positive vs 35% Tested, OR = 1.5 [1.3, 1.7]; p<0.001) and Inpatient admission (51% Inpatient vs 39% COVID-19 positive, OR = 1.7 [1.2, 2.4]; p=0.002). African American individuals had a significantly increased COVID-19 susceptibility risk (8% COVID-19 positive vs 7% Tested, OR = 1.3 [1.0, 1.6]; p=0.03) and a similar, non-significant increased risk for COVID-19 inpatient admission and severe outcome (11% Inpatient, 16% Severe) (Table 1). These findings show that self-identified non-white individuals had a higher risk of COVID-19 disease.

### Prior Diagnoses as COVID-19 Risk Factors

To identify diagnoses that were risk factors for COVID-19 severity groups, we analyzed diagnoses entered in the EHR *prior* to an individual’s SARS-CoV-2 testing. ICD codes were mapped to ∼1,800 phecodes, which have been shown to represent meaningful and interpretable phenotypes^20^. We sought COVID-19 susceptibility risk factors by comparing COVID-19 positive individuals to a random sampling of 10,000 individuals who were age- and sex-matched and drawn from the DDR cohort. In addition, we analyzed the presence of prior diagnoses between the COVID-19 positive Inpatient group compared to the COVID-19 positive Outpatient group and between the COVID-19 positive inpatient Severe group compared to COVID-19 positive inpatient not Severe group (Figure 1).

Three hundred eighty-six phecodes present prior to SARS-CoV-2 testing were associated with COVID-19 diagnosis susceptibility and included known risk factors such as cardiovascular disease, type 2 diabetes, hypercholesterolemia, hypertension, and chronic renal disease (Table S3) when adjusting for age and sex (Methods). To identify phenotypic risk factors not correlated with and not already explained by major known COVID-19 risk factors, we controlled for these major known COVID-19 risk factors (coronary heart disease, congestive heart failure, chronic obstructive pulmonary disease, type 2 diabetes, hyperlipidemia, hypertension, obesity, and chronic renal disease^7,8,10,13–15,27^). We identified 80 phecodes observed prior to SARS-CoV-2 testing that were associated with COVID-19 susceptibility (Figure 2, Table S4). Less well-established significant COVID-19 diagnosis susceptibility risk factors included dementias (dementia (OR 5.2 [3.2-8.3], p=2.6 × 10^−10^), Alzheimer’s disease (OR 7.7 [3.9-15], p=1.2 × 10^−8^)), mental health conditions (major depressive disorder (OR 2.1 [1.6-2.8], p=1.1 × 10^−6^), anxiety disorder (OR 1.7 [1.4, 2.0], p=5.3 × 10^−7^)), and vitamin D deficiency (OR 1.8 [1.4-2.2], p=5.7 × 10^−6^) (Figure 2, Table S4).

**Figure 2.**
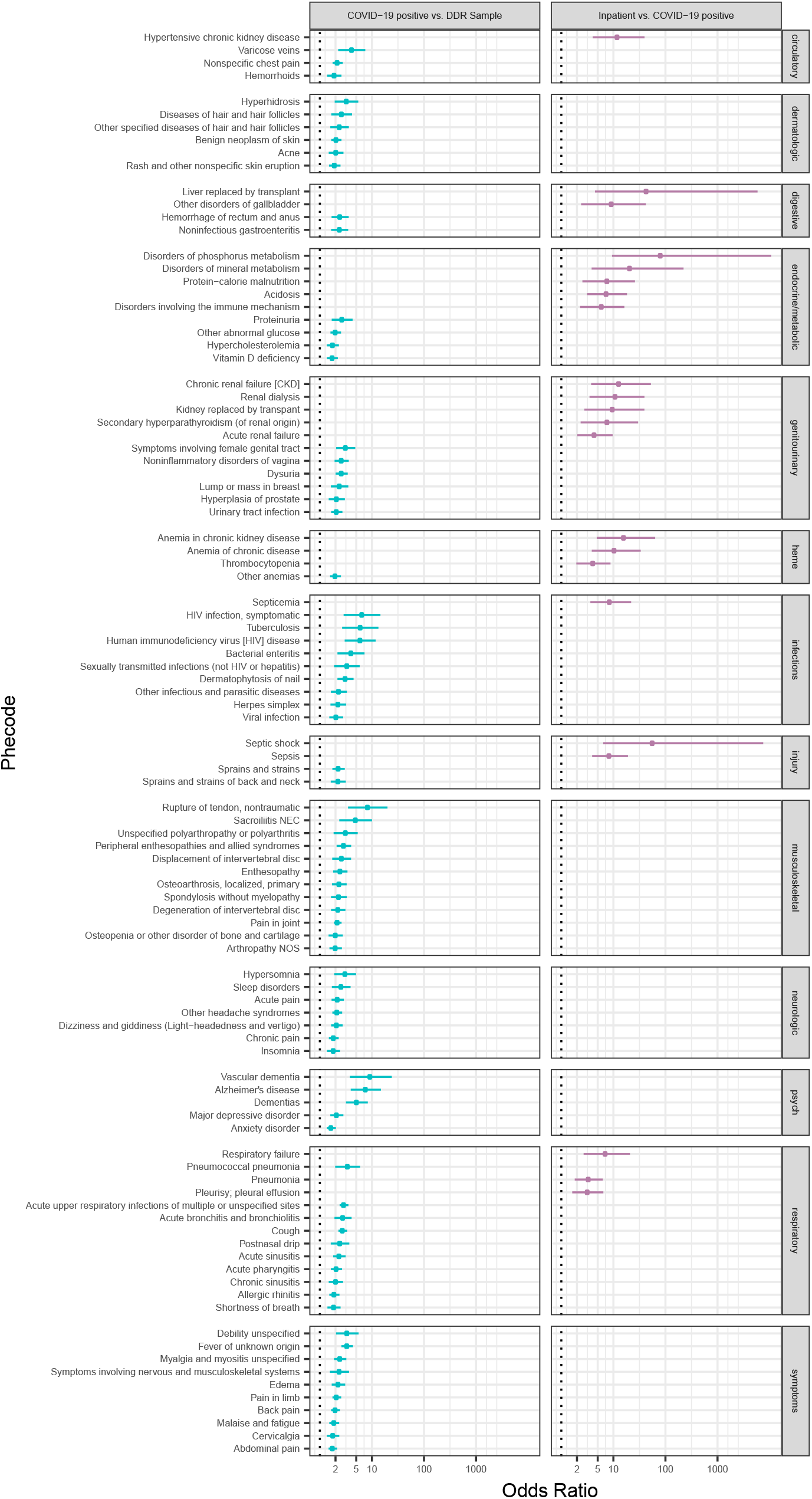
Significant prior phecode risk factors for COVID-19 susceptibility and inpatient correcting for age, sex and known risk factors (Model 2) grouped by phecode category. For phecode categories, dermatologic, congenital anomalies, and neoplasms were combined to form dermatologic. Neurological and sense organs were combined to form neurologic. heme = hematopoietic, psych = mental disorders.

Individuals with dementia and Alzheimer’s disease may be at greater risk of contracting COVID-19 as they are older and have more comorbidities^28^. In the DDR sample, patients over 65 years old with dementia have on average 2.3 known risk factors (standard deviation (SD) = 1.8) versus patients over 65 years old without dementia who have on average 0.98 known risk factors (SD = 1.4) (p<0.001). However, these comorbidities are less likely to confound these analyses, as we controlled for age and known comorbid risk factors. Individuals with dementia may also have limited access to accurate information and live in facilities that have high infection rates^28–32^.

Mental health conditions, including depression and anxiety, have not been described as prior risk factors for COVID-19 susceptibility. This may be due to the known association of mental health conditions and health outcomes including pneumonia and barriers to accessing health care^33,34^. We evaluated if these mental health associations were present in the UCLA Health system. Using the DDR Sample controlling for age and sex, we found that both depression and anxiety were positively associated with risk of influenza (depression OR 4.5 [2.6, 7.1], p<0.001; anxiety OR 2.2 [1.4, 3.2], p<0.001). Access to health care was likely not a contributing factor in our health system as patients with depression or anxiety compared to those without depression or anxiety had a greater number of health system encounters, after controlling for age and sex (20 encounters per year depression or anxiety vs 10 encounters per year without depression or anxiety, OR per 10 encounters per year 1.5 [1.4, 1.6], p<0.001).

The association of low vitamin D levels with COVID-19 disease has been proposed because rates of infection are higher in countries at higher latitudes^35–37^. To further investigate if vitamin D insufficiency was associated with COVID-19 susceptibility, we evaluated 25-hydroxyvitamin D laboratory values. Indeed, we found that the minimum level of 25-hydroxyvitamin D one year prior to SARS-CoV-2 testing was significantly lower in COVID-19 positive cases compared to the DDR sample, when correcting for age and sex (mean (SD) 29.1 ng/mL (11.7) COVID-19 positive vs 30.4 ng/mL (13.0), OR per 5 ng/mL 0.92 [0.84, 0.99], p=0.045), but not significant after correcting for age, sex and known risk factors (OR per 5 ng/mL 0.99 [0.88, 1.0], p=0.28) (Methods). These differences in vitamin D levels may not be clinically significant at an individual level, but support that vitamin D deficiency across the cohort increases the risk of COVID-19 diagnosis.

Additional significant prior risk factors associated with COVID-19 diagnosis susceptibility included phecodes representing common symptoms of COVID-19^8,9,15^. These included respiratory symptoms (cough (OR 2.7 [2.3, 3.3], p<2.2 × 10^−16^)), musculoskeletal symptoms (chronic pain (OR 1.9 [1.5, 2.3], p=7.2 × 10^−8^), and malaise and fatigue (OR 1.9 [1.5, 2.4], p=3.3 × 10^−8^)) (Figure 2, Table S4).

Phecodes present prior to SARS-CoV-2 testing significantly associated with the COVID-19 positive Inpatient group were renal disease (renal dialysis, acute renal failure), respiratory disease (pneumonia, pleurisy), and sepsis (Table S5). Controlling for known risk factors, 22 significant phecodes were identified as risk factors for the COVID-19 positive Inpatient group. Among these, less well-established risk factors included individuals with a history of acidosis (OR 7.4 [3.1, 18], 6.9×10^−6^), sepsis (OR 8.4 [3.9, 19], p=2.5×10^−8^), kidney replaced by renal transplant (OR 9.7 [2.8, 39], p=3.2×10^−4^), and disorders involving the immune mechanism (OR 6.0 [2.3, 16], p=2.7×10^−4^) (Figure 2, Table S6).

As acidosis and sepsis are typically diagnosed during relatively severe inpatient admissions, these factors may represent patients with a severe hospitalization prior to SARS-CoV-2 testing. Previous studies suggested a more rapid COVID-19 clinical progression in renal transplant patients^38–40^, but they did not investigate if renal transplant was associated with increased COVID-19 inpatient admission risk. Lastly, the “disorders involving the immune mechanism” phecode maps to ICD codes including miscellaneous immunodeficiency diseases (but not including humoral or T-cell defects). Immunodeficiency has yet to be shown as a COVID-19 risk factor for inpatient admission.

Even after controlling for chronic renal disease and other major known COVID-19 risk factors, multiple phenotypes associated with renal disease remained significantly associated with COVID-19 inpatient admission risk including kidney replaced by renal transplant as described above, acute renal failure (OR 4.4 [2.0, 9.7], p=1.5×10^−4^), renal dialysis (OR 11 [3.5, 39], p=2.5×10^−5^), and anemia in chronic kidney disease (OR 16 [4.8, 63], p=2.2×10^−6^) (Figure 2, Table S6).

A previous history of thrombocytopenia was also significantly associated with COVID-19 inpatient admission risk (OR 4.1 [1.9, 8.8], p=1.3×10^−4^). Many prior studies showed that thrombocytopenia during hospitalization is a risk factor for severe COVID-19 outcome^8,9,15,41,42^. We confirmed via laboratory test results that the minimum platelet level one year prior to SARS-CoV-2 testing was lower in COVID-19 inpatient cases compared to the COVID-19 outpatient cases, when correcting for sex, age and known risk factors (mean (SD) 173 ×10^3^/μL (82) COVID-19 Inpatient vs 240 ng/mL (66) COVID-19 Outpatient; OR per 10 ×10^3^/μL 0.89 [0.86, 0.93], p<0.001) (Methods).

We did not find any phecodes significantly associated with the COVID-19 Severe group, although our power was low (N=56 COVID-19 positive with Severe outcome and at least one encounter prior to SARS-CoV-2 testing) compared to the COVID-19 Inpatient group (N=177) and COVID-19 positive group (N=843).

### Prior Renal Diagnoses as COVID-19 Risk Factors

Given the strong association of additional renal diseases with COVID-19 risk of hospitalization even after controlling for chronic renal disease and other major known COVID-19 risk factors, we performed a detailed analysis of renal diseases. We investigated renal disease categories of: chronic renal disease, acute renal disease, end stage renal disease, anemia due to chronic renal disease, and renal transplant (See Table S7 for phecodes assigned to renal disease categories). Individuals with “any renal disease category” had at least one phecode corresponding the renal disease categories. We controlled for non-renal major known risk factors (coronary heart disease, congestive heart failure, chronic obstructive pulmonary disease, type 2 diabetes, hyperlipidemia, hypertension, and obesity) to determine the association of each renal disease category with COVID-19 diagnosis susceptibility and inpatient admission risk. As expected, all renal disease categories were significantly associated with COVID-19 inpatient risk. In addition, chronic renal disease, end-stage renal disease, anemia in chronic renal disease, and any renal disease category were significantly associated with COVID-19 diagnosis susceptibility, suggesting multiple renal diseases are risk factors for both COVID-19 diagnosis susceptibility and inpatient admission risk (Table 2).

**Table 2:** Association of prior renal diagnoses with COVID-19 diagnosis susceptibility and inpatient admission. DDR = DDR Sample, OR = odds ratio, CI = Confidence Interval, *p<0.05

| **Renal Diagnoses** | **COVID-19 positive (N=843) vs**<br>**DDR (N=10,000)** |  |  | **Inpatient (N=177) vs**<br>**Outpatient (N=666)** |  |  |
| --- | --- | --- | --- | --- | --- | --- |
|  | **OR [95% CI]** | **N<sub>COVID-19</sub>** | **N<sub>DDR</sub>** | **OR [95% CI]** | **N<sub>Inpatient</sub>** | **N<sub>Outpatient</sub>** |
| Chronic renal disease | 2.1 [1.5, 2.8]* | 89 | 241 | 2.3 [1.3, 4.3]* | 52 | 37 |
| Acute renal disease | 1.4 [0.94, 2.0] | 59 | 168 | 5.2 [2.5, 11]* | 43 | 16 |
| End-stage, stage V, dialysis | 2.0 [1.3, 3.1]* | 36 | 99 | 6.0 [2.5, 15]* | 27 | 9 |
| Anemia in chronic renal disease | 3.1 [1.8, 5.2]* | 32 | 42 | 17 [5.7, 59]* | 28 | 4 |
| Renal Transplant | 1.7 [0.86, 3.1] | 16 | 43 | 13 [4.1, 49]* | 12 | 4 |
| Any Renal Disease Category | 1.7 [1.3, 2.3]* | 105 | 351 | 2.8 [1.5, 5.0]* | 62 | 43 |

### Extremes of COVID-19 severity susceptibility or resistance

Next, we identified patients that were outliers based on what would be an expected COVID-19 clinical course predicated on their major predictive factors, namely age and diagnoses. These outlier groups included: 1) patients that were young with no major comorbidities (<25 years old), but who were admitted as an inpatient, and 2) older patients with high risk for a serious COVID-19 clinical course (>70 years old with at least three of the following: history of renal disease, diabetes, congestive heart failure, hyperlipidemia, and hypertension) with no inpatient admission. These patients may represent extremes of COVID-19 susceptibility and resistance, and therefore may carry genetic immunological susceptibilities or resistances to infection^43–46^. Out of the 77 young individuals with no major comorbidities, 7 (9%) were admitted to the hospital. Out of the 73 older individuals with a high risk for inpatient admission, 23 (30%) were not admitted to the hospital.

### Prior Medications Associated with COVID-19

Controversy remains over the protective or detrimental effects of medication classes including angiotensin converting enzyme inhibitors (ACEI), angiotensin receptor blockers (ARB)^47–49^, immunosuppressants^50–54^, steroids^55,56^, anticoagulants^57–59^, and non-steroidal anti-inflammatory drugs^60^. We investigated whether the use of these medications 90 days prior to SARS-CoV-2 testing (See Table S8 for drugs assigned to each medication class) was associated with COVID-19 susceptibility, inpatient admission, or severe outcome while controlling for age, sex and known risk factors (Methods). Comparing COVID-19 positive to negative individuals, those prescribed oral steroids had a decreased risk of being diagnosed with COVID-19 (OR 0.61 [0.45, 0.81], p<0.001). However, being prescribed oral steroids (OR 4.5 [2.3, 8.9], p<0.001) or other immunosuppressants (OR 4.1 [1.8, 9.0], p<0.001) increased the risk of inpatient admission. We did not observe that being prescribed ACEI or ARBs increased the risk of testing positive for SARS-CoV-2, being admitted to the hospital, or having a severe course, as recently reported^48^ (Figure 1, Table 3).

**Table 3.** Medications prior to SARS-CoV-2 testing associated with COVID-19 disease severity. OR = odds ratio, CI = confidence interval, pos=positive, neg=negative, Inp=Inpatient, Out = outpatient, Sev = Severe, not Sev = Not Severe, ACE inh = angiotensin converting enzyme inhibitors, ARBs = angiotensin receptor blockers, *p<0.05

|  | COVID-19 positive (N=843) vs<br>negative (N=23,790) |  |  | Inpatient (N=177) vs<br>Outpatient (N=666) |  |  | Severe (N=56) vs<br>not Severe (N=121) |  |  |
| --- | --- | --- | --- | --- | --- | --- | --- | --- | --- |
| Medication | OR [95% CI] | N<sub>pos</sub> | N<sub>neg</sub> | OR [95% CI] | N<sub>Inp</sub> | N<sub>Out</sub> | OR [95% CI] | N<sub>Sev</sub> | N<sub>not Sev</sub> |
| ACE Inh | 0.96 [0.63, 1.4] | 26 | 769 | 1.3 [0.53, 3.2] | 12 | 14 | 0.62 [0.14, 2.2] | 3 | 9 |
| ARBs | 1.2 [0.84, 1.7] | 39 | 994 | 1.1 [0.49, 2.5] | 15 | 24 | 0.50 [0.11, 1.8] | 3 | 12 |
| Immuno-suppressants | 0.90 [0.61, 1.3] | 32 | 994 | 4.1 [1.8, 9.0]* | 16 | 16 | 0.57 [0.14, 2.0] | 5 | 11 |
| Steroids | 0.61 [0.45, 0.81]* | 51 | 2303 | 4.5 [2.3, 8.9]* | 28 | 23 | 0.58 [0.2, 1.5] | 8 | 20 |
| Anticoagulant | 0.98 [0.59, 1.5] | 19 | 542 | 1.5 [0.48, 4.6] | 12 | 7 | 0.38 [0.04, 2.0] | 1 | 11 |
| NSAID | 0.86 [0.65, 1.1] | 58 | 1948 | 1.0 [0.48, 2.1] | 17 | 41 | 0.73 [0.22, 2.2] | 5 | 12 |

## Discussion

Identifying COVID-19 risk factors along the entire disease spectrum can inform patient care, public policies, and future research aimed at improving outcomes and reducing health care disparities. We leveraged our ability to query a de-identified electronic health records to determine COVID-19 risk factors in the UCLA Health System^61^. We found previously reported health disparities in the Hispanic or Latino, and non-white populations and confirmed known co-morbidities such as cardiovascular disease and chronic renal disease that are risk factors for COVID-19 susceptibility and inpatient admission. We also identified several less well-established risk factors such as pre-existing dementia, depression, vitamin D deficiency, and previous renal transplant. Furthermore, we found that immunosuppressive medication was associated with a decreased COVID-19 positive testing risk, but an increased inpatient admission risk.

This study differs from previous studies in that it was performed with direct EHR extraction as opposed to cases series, manual extraction, standardized form extraction from electronic health records, or registries^7,8,10,12,14,15,62^, allowing detailed phenotyping from extended historical data. Recent preprints have used direct EHR extraction from collaborations between the United States and South Korea^16^ and the United Kingdom^6^. The detailed phenotyping and increased power from these studies identified COVID-19 mortality risk factors in the United Kingdom not reported in manual EHR extraction studies including stroke, dementia, and autoimmune diseases (rheumatoid arthritis, lupus, psoriasis).

Significant risk factors for becoming critically ill (intensive care unit admission or death) among hospitalized COVID-19 patients include older age, cardiac disease (cardiac injury, coronary artery disease), pulmonary disease (chronic obstructive pulmonary disease, acute respiratory distress syndrome), renal disease (chronic renal disease, acute kidney injury), hypertension, diabetes, hyperlipidemia, and obesity^9–11,14,63,64^. Previously reported comorbid risk factors for severe outcome in COVID-19 hospitalized patients are also prior risk factors for COVID-19 diagnosis susceptibility and inpatient admission in our study. However, we find risk factors that are distinct to COVID-19 susceptibility and inpatient admission. For example, the risk of testing positive for COVID-19 increased with cardiovascular disease (coronary artery disease, congestive heart failure), hypertension, and hypercholesterolemia while risk factors for inpatient admission, were predominantly renal disorders (chronic renal failure, acute renal failure) and thrombocytopenia, the latter of which was confirmed via laboratory test results.

We identified prior risk factors that are less well-established for COVID-19 susceptibility including dementia, major depressive disorder, anxiety disorder and vitamin D deficiency. Individuals with dementia may be at greater risk of contracting COVID-19 as they are older and have more comorbidities, although we corrected for these in our analysis. We suspect that other environmental factors are contributing, including living in facilities which have had high infection rates, difficulty adhering to public health recommendations and limited access to accurate information^28–32^.

Studies on the mental health impact of COVID-19 have shown an increased rate of depressive symptoms^65,66^. Depression and anxiety as prior risk factors for COVID-19 susceptibility have not been previously described. Previous studies have correlated mental health condition with respiratory disease including pneumonia and pneumococcal disease^33^. We confirmed in the UCLA health system that individuals with depression and anxiety have an increased risk of influenza. These associations may be due to diminished effort for personal protection and barriers to accessing health care^33,34^, although we did not find that patients with depression or anxiety had less UCLA Health System encounters. Given that dementia and mental health conditions are risk factors for COVID-19 susceptibility, healthcare organizations and communities should consider strategies in addition to social distancing to reduce this risk, as social distancing may contribute to the risk^67^.

There has been speculation using country-wide aggregate data that low vitamin D may be associated with COVID-19, as rates of infection are higher in countries at higher latitudes^35– 37^. While there are many other explanations for this latitude effect (e.g. rates of testing, global patterns of spread), here, we show using individual patient data that vitamin D deficiency is associated with increased COVID-19 susceptibility risk. A recent preprint showed that low serum 25-hydroxycholecalcifoerol was more prevalent in ICU compared to non-ICU hospitalized patients^68^. We confirmed the association of vitamin D insufficiency with COVID-19 susceptibility using 25-hydroxyvitamin D laboratory levels when correcting for age and sex, but not when correcting for known risk factors. We did not evaluate whether vitamin D supplementation would be protective.

Less well-established inpatient admission prior risk factors included renal transplant and disorders involving the immune mechanism. Studies from Italy, New York, and China have reported the clinical course of hospitalized renal transplant patients with COVID-19 suggesting some may have a more rapid clinical progression^38–40^. Our study also shows that renal transplant patients have an increased risk of inpatient admission with COVID-19.

While there is controversy over whether immunosuppressant medication increases risk of COVID-19 severity^50–54^, we found that individuals with the “disorders involving the immune mechanism” phecode, which maps to ICD codes including immunodeficiency, were at increased risk of inpatient admission. Further studies are warranted to determine if other clinical characteristics of this patient group increase the risk of COVID-19 severity and if there are mechanistic links.

In our study cohort, older age was not a risk factor for all stages of the COVID-19 disease spectrum. Individuals older than 65 years old were more likely to be admitted, but made up a lower percentage of individuals experiencing a severe outcome. As a corollary, young age was not completely protective, as those in the intermediate age group between 26 and 55 years old were more likely to have a severe outcome once admitted. Studies from the CDC and New York showed intensive care unit admission or ventilation was lower in intermediate age groups compared to older age groups^69–71^, in contrast to our study. In the UCLA Health System, it is possible clinicians have a lower threshold of inpatient admission for older individuals, preferring to monitor their COVID-19 clinical course. However, less of these admitted older individuals progress to severe disease. Clinicians may have a higher threshold for admitting intermediate age group individuals where those admitted may have more significant COVID-19 symptoms. Therefore, once admitted intermediate age group individuals are more likely to progress to severe disease. Risk factors other than age and co-morbidities likely also contribute to COVID-19 severe outcomes. These risk factors may be environmental such as air pollution^72^, socioeconomic such as access to health care^26,73^, and genetic^43,74^.

The increased COVID-19 disease susceptibility and severity risk in individuals with self-identified Hispanic or Latino ethnicity further highlights health disparities in COVID-19. We found a similar association in individuals with self-identified race other than White or Caucasian and self-identified Black or African American race. The Los Angeles County of Public Health has confirmed these disparities; the age-adjusted rate of COVID-19 cases is over 100 per 100,000 individuals for these minority groups but only 78 for those who self-identify as white, non-hispanic^22^. Similar findings have been found by the New York City Health Department and Chicago Department of Public health where the rates of COVID-19 cases and severe outcomes are roughly double in Black and Hispanic or Latino individuals compared to White individuals^21,23^. Other studies from Connecticut and a 14 state public health COVID-19 surveillance network (COVID-NET) including California predominantly found risk of COVID-19 susceptibility and hospitalization in Black individuals^70,75^. As we are unable to appropriately assess socioeconomic status using the available EHR data, ethnic discrepancy may be due to this and other associated variables^76^. For example, shelter-in-place guidelines are most burdensome to Hispanic or Latino individuals in Los Angeles County as many live in neighborhoods with densely populated communities and have limited access to both open spaces and nearby supermarkets^26^.

There has been concern that some individuals with COVID-19 have an exaggerated immune response resulting in cytokine storm or secondary haemophagocytic lymphohistiocytosis, and studies have suggested that short-term immunosuppression is warranted^51,54,77^. Previous studies investigated small numbers of COVID-19 positive patients (<4) on immunosuppression prior to COVID-19 diagnosis, finding that these patients in general did well and did not have a severe outcome^50,52,53^. Here we analyzed over 80 patients on either oral steroids or other immunosuppressants. Individuals on oral steroids had a decreased COVID-19 positive testing rate, but an increased inpatient admission risk. The explanation for this may be biological, behavioral, or partially due to clinician bias. It is possible immunosuppression results in asymptomatic COVID-19, thus lowering numbers of immunosuppressed individuals who seek testing and therefore test positive. However, if positive, these individuals have severe symptoms requiring admission. Another explanation is that clinicians lower their threshold for inpatient admission for patients on immunosuppression. We did not assess whether immunosuppressive medication was stopped, continued or changed on admission, and therefore, we are unable to draw conclusions on whether to adjust immunosuppression in COVID-19 hospitalized patients.

This study identifies important and less established risk factors for susceptibility and severity of outcomes in COVID-19. These risk factors should spur future work in understanding the mechanistic underpinnings of these observations and implementing strategies to mitigate both biological and socioeconomic risk factors.

## Data Availability

The EHR dataset is not publicly available due to reasonable privacy and security concerns unless researchers are among the Institutional Review Board-approved research collaborations with the named medical centers.

## UCLA Health Data Mart Working Group

The UCLA Health Data Mart Working Group is comprised of the Computational Medicine Data Mart Working Group: Brunilda Balliu, Ph.D.; Yael Berkovich, B.A., Eleazar Eskin, Ph.D., Eran Halperin, Ph.D., Brian L. Hill, M.S., Nadav Rakocz, B.Sc, Akos Rudas, M.Sc; the Office of Health Informatics and Analytics Data Mart Working Group: Anna Liza Malazarte Antonio, DrPH, Angela Marquessa Badillo, R.N., Michael Broudy, B.A., Tony Dang, Ankur Jain, M.S., Vivek Katakwar, B.Sc., Sheila Minton, Ghouse Mohammed, B.Tech, Ariff Muhamed, Pabba Pavan, Michael A. Pfeffer, M.D., Rey Salonga, Timothy J Sanders, B.A., Paul Tung, B.S., Vu Vu, B.Sc., Ailsa Zheng, B.A; and the Institute for Precision Health Data Mart Working Group: Maryam Ariannejad, Chris Denny, M.D., Clara Lajonchere, Ph.D., Clara Magyar, Ph.D

## Author Contributions

Conceptualization, T.S.C, Y.D., M.K.F., R.J., T.S., J.M.Y., C.H., A.W., D.H.G., M.J.B., and B.P.; Methodology, T.S.C, Y.D., M.K.F., J.M.Y., C.H., M.J.B., and B.P.; Software, T.S.C, Y.D., M.K.F., R.J., and T.S.; Validation, T.S.C, Y.D., M.K.F., R.J., and T.S.; Formal Analysis, T.S.C, Y.D., M.K.F., R.J., and T.S.; Investigation, T.S.C, Y.D., M.K.F., R.J., and T.S.; Resources, D.H.G., M.J.B., B.P., and UCLA Precision Health Data Discovery Repository Working Group; Data Curation, T.S.C, Y.D., M.K.F., R.J., T.S., and UCLA Precision Health Data Discovery Repository Working Group; Writing – Original Draft, T.S.C, M.K.F., D.H.G., M.J.B., and B.P.; Writing – Review & Editing, T.S.C, Y.D., M.K.F., R.J., T.S., J.M.Y., C.H., D.H.G., M.J.B., and B.P.; Visualization, T.S.C, Y.D., and M.K.F.; Supervision, T.S.C., M.J.B., and B.P.; Funding Acquisition, D.H.G., M.J.B., and B.P.

## Methods

### Study design

This was an observational case-control study within a cohort of patients registered at the UCLA Health System after January 1, 2013. The UCLA Health System includes two hospitals (520 and 281 inpatient beds) and 210 primary and specialty outpatient locations predominantly located in Los Angeles County. We leveraged an extract of the de-identified EHR from the UCLA Health System known as the UCLA Data Discovery Repository (DDR), developed under the auspices of the UCLA Health Office of Health Informatics Analytics and the UCLA Institute of Precision Health. The DDR contains longitudinal electronic records for more than 1.5 million patients since 2013, including patient demographics, problems, medications, vital signs, past medical history, and laboratory data. This study was considered human subjects research exempt because all electronic health records were de-identified (UCLA IRB# 20-001180).

### Outcomes and Case/Control Selection

From March 9, 2020 to June 14, 2020, 26,602 individuals were tested for SARS-CoV-2 via reverse transcriptase polymerase chain reaction (RT-PCR) within the UCLA Health System. Three different levels of COVID-19 outcome were defined (Figure 1).

COVID-19 positive cases were those with a positive SARS-CoV-2 PCR test result (COVID-19 positive). To determine individuals with SARS-CoV-2 testing, lab tests in the DDR were searched for “SARS-CoV-2” or “COVID-19”. The list of tests was manually reviewed to determine which tests were PCR based. SARS-CoV-2 PCR tests in our hospital system reported positive tests as “positive” or “detected” and negative tests as “negative” or “not detected”. Other lab values such as “unknown” or “NULL” were considered unknown. Some individuals had more than one SARS-CoV-2 PCR test; in these cases, individuals were considered COVID-19 positive if they had at least one positive test. The controls for COVID-19 positive cases were 10,000 random individuals from the 1.5 million DDR patients without a SARS-CoV-2 PCR test that were sex and age group matched to COVID-19 positive individuals (DDR Sample). To study whether medications taken prior to COVID-19 testing were associated with COVID-19 diagnosis, we examined the 90-day window prior to the test. This window necessitated comparison to COVID-19 negative individuals as controls (COVID-19 negative).

Inpatient cases were those who had a hospital admission within 14 days of their first positive SARS-CoV-2 PCR test (Inpatients). This time window was selected to identify individuals whose symptoms may not have been severe at initial testing, but may have progressed in severity warranting inpatient admission. The controls for inpatient cases were those who tested positive for COVD-19, but were not admitted as inpatient within 14 days of their first positive SARS-CoV-2 PCR test (Outpatients).

Severe cases were those who were admitted to an intensive care unit or were intubated within 14 days of their first positive SARS-CoV-2 PCR test (Severe). Intubation was determined by CPT codes (94656, 94657, 94662, 1120200004, 1120200012) and codes custom to the UCLA EHR (Table S9). The controls for Severe cases were those who had a hospital admission within 14 days of their first positive SARS-CoV-2 PCR test, but were not admitted to an intensive care unit and not intubated during the 14-day window (Inpatient Not Severe).

For prior diagnoses, lab and medication association test, cases and controls were excluded if their records were missing age, missing sex, or had no encounter in the UCLA Health System prior to the SARS-CoV-2 PCR test.

### Confounding variables

We treated sex, age, age^2 and prior diagnoses of major known COVID-19 risk factors (coronary heart disease, congestive heart failure, chronic obstructive pulmonary disease, type 2 diabetes, hyperlipidemia, hypertension, obesity, and chronic renal disease^7,8,10,13–15^) as potential confounders and adjusted for all or a subset of them depending on the analyses (see Table S1 for phecodes assigned to risk factors). For all analyses using 10,000 DDR individuals, the matching factor age group was also included as a potential confounder.

### Statistical Analysis

#### Demographics

Age, sex, race and ethnicity were stratified by the Tested, COVID-19 positive, Inpatient and Severe groups. For age groups and sex, percentages of Severe was compared to Inpatient, Inpatient was compared to COVID-19 positive, and COVID-19 positive was compared to Tested using a Fisher’s exact test^78^. Age groups were: <18 years, 19-25 years, 26-55 years, 56-65 years, and >65 years. Significant association of race and ethnicity in a COVID-19 outcome was analyzed in the following Firth logistic regression models^79^.

~~~
logit(outcome)=*β*_*0*_ + *β*_*1*_race + *β_2_*age_group + *β*_*3*_sex
~~~

and

~~~
logit(outcome)= *β*_*0*_ + *β*_*1*_ethnicity + *β*_*2*_age_group + *β*_*3*_sex
~~~

Outcome for Severe was considered Inpatient Severe vs. Inpatient Not Severe; for Inpatient was considered COVID-19 positive Inpatient vs COVID-19 positive Outpatient; and for COVID-19 positive was considered COVID-19 positive vs. COVID-19 negative. Individuals with unknown race and unknown ethnicity were excluded from these analyses.

For Hispanic or Latino ethnicity, we also corrected for patient histories of known risk factors (coronary heart disease, congestive heart failure, chronic obstructive pulmonary disease, type 2 diabetes, hyperlipidemia, hypertension, obesity, and chronic renal disease) (see Table S1 for phecodes assigned to risk factors) using the model:

~~~
logit(outcome)=*β*_*0*_ + *β*_*1*_Hispanic or Latino ethnicity + *β*_*2*_age_group + *β*_*3*_sex + *β*_*4*_coronary_artery_disease + *β*_*5*_congestive_heart_failure + *β*_*6*_chronic_obstructive_pulmonary_disease + *β*_*7*_type_2_diabetes + *β*_*8*_hyperlipidemia + *β*_*9*_hypertension + *β*_*10*_obesity + *β*_*11*_chronic_renal_disease
~~~

### Prior Diagnoses as COVID-19 Risk Factors

International Statistical Classification of Diseases (ICD)-9-CM and ICD-10-CM codes were mapped to 1866 phecodes, which represent meaningful and interpretable phenotypes^20,80^. We calculated the odds ratio of all phecodes correcting for age and sex (Model 1) using the Firth logistic regression model^81,82^ to account for phecodes with no counts within an outcome group as follows:

~~~
(Model 1) logit(outcome)=*β*_*0*_ + *β*_*1*_age + *β*_*2*_age^2^ + *β*_*3*_sex + *β*_*4*_phecode
~~~

Outcome for Severe was considered Inpatient Severe vs. Inpatient Not Severe; for Inpatient was considered COVID-19 positive Inpatient vs COVID-19 positive Outpatient; and for COVID-19 positive was considered COVID-19 positive vs. the DDR Sample.

For COVID-19 positive compared to the DDR Sample, we added an age group covariate. Age group, age and age squared are included in the model to correct for residual confounding effects of age due to possible non-linear effect. We used the likelihood ratio test to test for phecode significance^79^. Multiple hypothesis testing correction was performed conservatively using a Bonferroni correction for the number of phecodes tested^83^. Nominal p-values are reported.

To account for major known risk factors (coronary heart disease, congestive heart failure, chronic obstructive pulmonary disease, type 2 diabetes, hyperlipidemia, hypertension, obesity, and chronic renal disease), we tested Bonferroni significant phecodes from Model 1 in Model 2 using the Firth logistic regression model as follows:

~~~
(Model 2) logit(outcome)=*β*_*0*_ + *β*_*1*_age + *β*_*2*_age^2^ + *β*_*3*_sex + *β*_*4*_coronary_artery_disease + *β*_*5*_congestive_heart_failure + *β*_*6*_chronic_obstructive_pulmonary_disease + *β*_*7*_type_2_diabetes + *β*_*8*_hyperlipidemia + *β*_*9*_hypertension + *β*_*10*_obesity + *β*_*11*_chronic_renal_disease + *β*_*12*_phecode
~~~

For COVID-19 positive compared to the DDR Sample, we added an age group covariate. Bonferroni correction was performed for the number of phecodes tested in Model 2.

We performed secondary analyses for significant risk factors associated with COVID-19 susceptibility and inpatient admission. Because dementia and Alzheimer’s disease were associated with COVID-19 susceptibility, we determined if dementia patients had more known risk factors. From the DDR sample, we calculated the mean number of known risk factors for patients over 65 years old with the dementia phecode (290.1) and patients over 65 years old without the dementia phecode. We compared these groups using the Wilcoxan rank sum test.

Because the mental health conditions, depression and anxiety, were associated with COVID-19 susceptibility, we determined if previously described explanations may account for this association. We determined if depression (phecode 296.22) and anxiety (phecode 300.10) were associated with influenza (phecode 418.00) in the DDR Sample in following Firth logistic regression model and used the likelihood ratio test:

~~~
logit(influenza)=*β*_*0*_ + *β*_*1*_age + *β*_*2*_age^2^ + *β*_*4*_age_group + *β*_*4*_sex + *β*_*5*_depression
~~~

and

~~~
logit(influenza)=*β*_*0*_ + *β*_*1*_age + *β*_*2*_age^2^ + *β*_*4*_age_group + *β*_*4*_sex + *β*_*5*_anxiety
~~~

As a proxy for access to health care, we determined if the number of encounters per year was associated with depression or anxiety in the DDR Sample. We considered encounters after 2013 as this was when the EHR was implemented. The total number of encounters for an individual was defined as the number of encounters with unique dates. This prevented counting multiple encounters on the same date more than once. Individuals were included if the first and last encounter in the EHR was at least 6 months apart. 5613 of the 10,000 individuals in the DDR Sample met these criteria. We tested the association of number of encounters per year with depression or anxiety in the following Firth logistic regression model and used the likelihood ratio test:

~~~
logit(depression or anxiety)=*β*_*0*_ + *β*_*1*_age + *β*_*2*_age^2^ + *β*_*4*_age_group + *β*_*4*_sex + *β*_*5*_encounters_per_year
~~~

We evaluated laboratory tests for laboratory-related phecodes associated with COVID-19 susceptibility, which included Vitamin D insufficiency, and inpatient admission, which included thrombocytopenia. For vitamin D insufficiency, we identified the minimum 25-hydroxyvitamin D within the past one year of SARS-CoV-2 testing. We reasoned the minimum value would more accurately reflect the level of vitamin D insufficiency as higher values may result from vitamin D supplementation. We compared the minimum 25-hydroxyvitamin D in COVID-19 positive (measured in 138 of 843) to the DDR Sample (measured in 487 of 10,000) with the following Firth logistic regression model and used the likelihood ratio test:

~~~
logit(outcome)=*β*_*0*_ + *β*_*1*_age + *β*_*2*_age_group + *β*_*3*_age^2^ + *β*_*4*_sex + *β*_*5*_25-hydroxyvitamin_D
~~~

and

~~~
logit(outcome)=*β*_*0*_ + *β*_*1*_age + *β*_*2*_age_group + *β*_*3*_age^2^ + *β*_*4*_sex + *β*_*5*_coronary_artery_disease + *β*_*6*_congestive_heart_failure + *β*_*7*_chronic_obstructive_pulmonary_disease + *β*_*8*_type_2_diabetes + *β*_*9*_hyperlipidemia + *β*_*10*_hypertension + *β*_*11*_obesity + *β*_*12*_chronic_renal_disease + *β*_*13*_25-hydroxyvitamin_D
~~~

For thrombocytopenia, we identified the minimum platelet level within the past one year of SARS-CoV-2 testing. We reasoned the minimum value would more accurately reflect when a diagnosis of thrombocytopenia would be made. We compared the minimum platelet level in COVID-19 inpatients (measured in 105 of 177) to the COVID-19 outpatients (measured in 277 of 666) with the following Firth logistic regression model and used the likelihood ratio test:

~~~
logit(outcome)=*β*_*0*_ + *β*_*1*_age + *β*_*2*_age^2^ + *β*_*3*_sex + *β*_*4*_coronary_artery_disease + *β*_*5*_congestive_heart_failure + *β*_*6*_chronic_obstructive_pulmonary_disease + *β*_*7*_type_2_diabetes + *β*_*8*_hyperlipidemia + *β*_*9*_hypertension + *β*_*10*_obesity + *β*_*11*_chronic_renal_disease + *β*_*12*_platelet
~~~

### Prior Renal Diagnoses as COVID-19 Risk Factors

We considered the following renal disease categories: chronic renal disease, acute renal disease, end stage renal disease, anemia due to chronic renal disease, and renal transplant (see Table S7 for phecodes assigned to renal disease categories). We controlled for major known risk factors except for chronic renal disease (coronary heart disease, congestive heart failure, chronic obstructive pulmonary disease, type 2 diabetes, hyperlipidemia, hypertension, and obesity) (Table S1) in the following Firth logistic regression model and used the likelihood ratio test:

~~~
logit(outcome)= *β*_*0*_ + *β*_*1*_age+ *β*_*2*_age^2^ + *β*_*3*_sex + *β*_*4*_coronary_artery_disease + *β*_*5*_congestive_heart_failure + *β*_*6*_chronic_obstructive_pulmonary_disease + *β*_*7*_type_2_diabetes + *β*_*8*_hyperlipidemia + *β*_*9*_hypertension + *β*_*10*_obesity + *β*_*11*_renal_category
~~~

Outcome for Severe was considered Inpatient Severe vs. Inpatient Not Severe; for Inpatient was considered COVID-19 positive Inpatient vs COVID-19 positive Outpatient; and for COVID-19 positive was considered COVID-19 positive vs. the DDR Sample.

### Extremes of COVID-19 severity susceptibility or resistance

We identified COVID-19 young patients with no major comorbidities admitted as an inpatient and COVID-19 older patients with comorbidities not admitted as an inpatient. We defined COVID-19 young patients with no major comorbidities admitted as an inpatient as:

- COVID-19 positive
- Less than 25 years old
- At least one prior UCLA Health encounter
- None of the following comorbidities: history of renal disease, diabetes, congestive heart failure, hyperlipidemia, and hypertension. These comorbidities were based on phecode groupings in Table S1.
- Inpatient admission within 14 days of a SARS-CoV-2 positive test.

We defined COVID-19 older patients with major comorbidities not admitted as an inpatient as:

- COVID-19 positive
- Greater than 70 years old
- At least one prior UCLA Health encounter
- At least three of the following comorbidities: history of renal disease, diabetes, congestive heart failure, hyperlipidemia, and hypertension. These comorbidities were based on phecode groupings in Table S1.
- No inpatient admission within 14 days of a SARS-CoV-2 positive test

### Prior Medications Associated with COVID-19

We investigated the use of angiotensin converting enzyme inhibitors (ACEI), angiotensin receptor blockers (ARB), immunosuppressants, oral steroids, oral anticoagulants, and non-steroidal anti-inflammatory drugs (see Table S8 for drugs assigned to medication classes) 90 days prior to SARS-CoV-2 testing. We controlled for age, sex, and known risk factors in the following Firth logistic regression model and used the likelihood ratio test:

~~~
logit(outcome)= *β*_*0*_ + *β*_*1*_age + *β*_*2*_age^2^ *β*_*3*_sex + *β*_*4*_coronary_artery_disease + *β*_*5*_congestive_heart_failure + *β*_*6*_chronic_obstructive_pulmonary_disease + *β*_*7*_type_2_diabetes + *β*_*8*_hyperlipidemia + *β*_*9*_hypertension + *β*_*10*_obesity + *β*_*11*_chronic_renal_disease + *β*_*12*_medication
~~~

Outcome for Severe was considered Inpatient Severe vs. Inpatient Not Severe; for Inpatient was considered COVID-19 positive Inpatient vs COVID-19 positive Outpatient; and for COVID-19 positive was considered COVID-19 positive vs. COVID-19 negative. We performed the latter comparison to preserve the 90-day prior medication usage window.

## Supplemental Information

Table S1. Known risk factor diagnoses and assigned phecodes

Table S3. Bonferroni corrected significant prior phecode risk factors for COVID-19 susceptibility correcting for age and sex (Model 1). Orange shaded cells indicate Bonferroni corrected significant phecodes where nominal p-value < Bonferroni critical p-value. Red shaded cells indicate phecodes with Bonferroni critical p-value < nominal p-value < 0.05. Unshaded cells indicate phecodes with nominal p-value > 0.05. Nominal p-values are reported. Phecodes not assigned to any patients or resulting in a model error were excluded.

Table S4. Bonferroni corrected significant prior phecode risk factors for COVID-19 susceptibility correcting for age, sex, and known risk factors (Model 2). Orange shaded cells indicate Bonferroni corrected significant phecodes where nominal p-value < Bonferroni critical p-value. Red shaded cells indicate phecodes with Bonferroni critical p-value < nominal p-value < 0.05. Unshaded cells indicate phecodes with nominal p-value > 0.05. Nominal p-values are reported. Phecodes not assigned to any patients or resulting in a model error were excluded.

Table S5. Bonferroni corrected significant prior phecode risk factors for COVID-19 inpatient admission correcting for age and sex (Model 1). Orange shaded cells indicate Bonferroni corrected significant phecodes where nominal p-value < Bonferroni critical p-value. Red shaded cells indicate phecodes with Bonferroni critical p-value < nominal p-value < 0.05. Unshaded cells indicate phecodes with nominal p-value > 0.05. Nominal p-values are reported. Phecodes not assigned to any patients or resulting in a model error were excluded.

Table S6. Bonferroni corrected significant prior phecode risk factors for COVID-19 inpatient admission correcting for age, sex, and known risk factors (Model 2). Orange shaded cells indicate Bonferroni corrected significant phecodes where nominal p-value < Bonferroni critical p-value. Red shaded cells indicate phecodes with Bonferroni critical p-value < nominal p-value < 0.05. Unshaded cells indicate phecodes with nominal p-value > 0.05. Nominal p-values are reported. Phecodes not assigned to any patients or resulting in a model error were excluded.

Table S8. Medication class and medication. ACE inhibitor = angiotensin converting enzyme inhibitors, ARBs = angiotensin receptor blockers.

## References

1. Dong, E., Du, H., and Gardner, L. (2020). An interactive web-based dashboard to track COVID-19 in real time. Lancet Infect Dis 20, 533–534.

2. Center for Systems Science and Engineering at Johns Hopkins University COVID-19 Map. Johns Hopkins Coronavirus Resource Center. https://coronavirus.jhu.edu/map.html.

3. Centers for Disease Control and Prevention (2020). CDC Activities and Initiatives Supporting the COVID-19 Response and the President’s Plan for Opening America Up Again.

4. The Lancet (2020). Sustaining containment of COVID-19 in China. Lancet 395, 1230.

5. Bhatraju, P.K., Ghassemieh, B.J., Nichols, M., Kim, R., Jerome, K.R., Nalla, A.K., Greninger, A.L., Pipavath, S., Wurfel, M.M., Evans, L., et al. (2020). Covid-19 in Critically Ill Patients in the Seattle Region - Case Series. N. Engl. J. Med. 382, 2012–2022.

6. Collaborative, T.O., Williamson, E., Walker, A.J., Bhaskaran, K.J., Bacon, S., Bates, C., Morton, C.E., Curtis, H.J., Mehrkar, A., Evans, D., et al. (2020). OpenSAFELY: factors associated with COVID-19-related hospital death in the linked electronic health records of 17 million adult NHS patients. medRxiv, 2020.05.06.20092999.

7. Grasselli, G., Zangrillo, A., Zanella, A., Antonelli, M., Cabrini, L., Castelli, A., Cereda, D., Coluccello, A., Foti, G., Fumagalli, R., et al. (2020). Baseline Characteristics and Outcomes of 1591 Patients Infected With SARS-CoV-2 Admitted to ICUs of the Lombardy Region, Italy. JAMA 323, 1574–1581.

8. Guan, W., Ni, Z., Hu, Y., Liang, W., Ou, C., He, J., Liu, L., Shan, H., Lei, C., Hui, D.S.C., et al. (2020). Clinical Characteristics of Coronavirus Disease 2019 in China. N Engl J Med.

9. Huang, C., Wang, Y., Li, X., Ren, L., Zhao, J., Hu, Y., Zhang, L., Fan, G., Xu, J., Gu, X., et al. (2020). Clinical features of patients infected with 2019 novel coronavirus in Wuhan, China. Lancet 395, 497–506.

10. Li, X., Xu, S., Yu, M., Wang, K., Tao, Y., Zhou, Y., Shi, J., Zhou, M., Wu, B., Yang, Z., et al. (2020). Risk factors for severity and mortality in adult COVID-19 inpatients in Wuhan. J Allergy Clin Immunol.

11. Lighter, J., Phillips, M., Hochman, S., Sterling, S., Johnson, D., Francois, F., and Stachel, A. (2020). Obesity in patients younger than 60 years is a risk factor for Covid-19 hospital admission. Clin Infect Dis.

12. Wu, Z., and McGoogan, J.M. (2020). Characteristics of and Important Lessons From the Coronavirus Disease 2019 (COVID-19) Outbreak in China: Summary of a Report of 72L314 Cases From the Chinese Center for Disease Control and Prevention. JAMA.

13. Yang, J., Zheng, Y., Gou, X., Pu, K., Chen, Z., Guo, Q., Ji, R., Wang, H., Wang, Y., and Zhou, Y. (2020). Prevalence of comorbidities and its effects in coronavirus disease 2019 patients: A systematic review and meta-analysis. Int. J. Infect. Dis. 94, 91–95.

14. Zhou, F., Yu, T., Du, R., Fan, G., Liu, Y., Liu, Z., Xiang, J., Wang, Y., Song, B., Gu, X., et al. (2020). Clinical course and risk factors for mortality of adult inpatients with COVID-19 in Wuhan, China: a retrospective cohort study. The Lancet 395, 1054–1062.

15. Goyal, P., Choi, J.J., Pinheiro, L.C., Schenck, E.J., Chen, R., Jabri, A., Satlin, M.J., Campion, T.R., Nahid, M., Ringel, J.B., et al. (2020). Clinical Characteristics of Covid-19 in New York City. N. Engl. J. Med.

16. Burn, E., You, S.C., Sena, A., Kostka, K., Abedtash, H., Abrahao, M.T.F., Alberga, A., Alghoul, H., Alser, O., Alshammari, T.M., et al. (2020). An international characterisation of patients hospitalised with COVID-19 and a comparison with those previously hospitalised with influenza. medRxiv, 2020.04.22.20074336.

17. Jordan, R.E., Adab, P., and Cheng, K.K. (2020). Covid-19: risk factors for severe disease and death. BMJ 368.

18. CDC (2020). Coronavirus Disease 2019 (COVID-19): Cases and Deaths by County. Centers for Disease Control and Prevention. https://www.cdc.gov/coronavirus/2019-ncov/cases-updates/county-map.html.

19. U.S. Census Bureau (2018). U.S. Census Bureau QuickFacts: Los Angeles County, California. https://www.census.gov/quickfacts/losangelescountycalifornia.

20. Wei, W.-Q., Bastarache, L.A., Carroll, R.J., Marlo, J.E., Osterman, T.J., Gamazon, E.R., Cox, N.J., Roden, D.M., and Denny, J.C. (2017). Evaluating phecodes, clinical classification software, and ICD-9-CM codes for phenome-wide association studies in the electronic health record. PLoS ONE 12, e0175508.

21. Chicago Department of Public Health (2020). Latest Data. https://www.chicago.gov/content/city/en/sites/covid-19/home/latest-data.html.

22. Los Angeles County Department of Public Health, Chief Science Office (2020). COVID-19 Racial, Ethnic & Socioeconomic Data & Strategies Report.

23. New York City Health (2020). Age-adjusted rates of lab confirmed COVID-19.

24. Dominguez, K., Penman-Aguilar, A., Chang, M.-H., Moonesinghe, R., Castellanos, T., Rodriguez-Lainz, A., and Schieber, R. (2015). Vital Signs: Leading Causes of Death, Prevalence of Diseases and Risk Factors, and Use of Health Services Among Hispanics in the United States — 2009–2013. MMWR Morb Mortal Wkly Rep 64, 469–478.

25. Velasco-Mondragon, E., Jimenez, A., Palladino-Davis, A.G., Davis, D., and Escamilla-Cejudo, J.A. (2016). Hispanic health in the USA: a scoping review of the literature. Public Health Reviews 37, 31.

26. Ong, P., Gonzalez, S., Pech, C., Diaz, S., Ong, J., Ong, E., and Aguilar, J. (2020). Struggling to Stay Home: How COVID-19 Shelter in Place Polocies Affect Los Angeles County’s Black and Latino Neighborhoods (UCLA Latino Policy and Politics Initiative, the UCLA Center for Neighborhood Knowledge, Ong & Associates).

27. Zhu, L., She, Z.-G., Cheng, X., Qin, J.-J., Zhang, X.-J., Cai, J., Lei, F., Wang, H., Xie, J., Wang, W., et al. (2020). Association of Blood Glucose Control and Outcomes in Patients with COVID-19 and Pre-existing Type 2 Diabetes. Cell Metab. 31, 1068-1077.e3.

28. Brown, E.E., Kumar, S., Rajji, T.K., Pollock, B.G., and Mulsant, B.H. (2020). Anticipating and Mitigating the Impact of the COVID-19 Pandemic on Alzheimer’s Disease and Related Dementias. Am J Geriatr Psychiatry.

29. McMichael, T.M., Currie, D.W., Clark, S., Pogosjans, S., Kay, M., Schwartz, N.G., Lewis, J., Baer, A., Kawakami, V., Lukoff, M.D., et al. (2020). Epidemiology of Covid-19 in a Long-Term Care Facility in King County, Washington. N. Engl. J. Med. 382, 2005–2011.

30. Onder, G., Rezza, G., and Brusaferro, S. (2020). Case-Fatality Rate and Characteristics of Patients Dying in Relation to COVID-19 in Italy. JAMA.

31. Wang, H., Li, T., Barbarino, P., Gauthier, S., Brodaty, H., Molinuevo, J.L., Xie, H., Sun, Y., Yu, E., Tang, Y., et al. (2020). Dementia care during COVID-19. Lancet 395, 1190–1191.

32. Roxby, A.C., Greninger, A.L., Hatfield, K.M., Lynch, J.B., Dellit, T.H., James, A., Taylor, J., Page, L.C., Kimball, A., Arons, M., et al. (2020). Outbreak Investigation of COVID-19 Among Residents and Staff of an Independent and Assisted Living Community for Older Adults in Seattle, Washington. JAMA Intern Med.

33. Seminog, O.O., and Goldacre, M.J. (2013). Risk of pneumonia and pneumococcal disease in people with severe mental illness: English record linkage studies. Thorax 68, 171–176.

34. Yao, H., Chen, J.-H., and Xu, Y.-F. (2020). Patients with mental health disorders in the COVID-19 epidemic. Lancet Psychiatry 7, e21.

35. Ilie, P.C., Stefanescu, S., and Smith, L. (2020). The role of vitamin D in the prevention of coronavirus disease 2019 infection and mortality. Aging Clin Exp Res.

36. Rhodes, J.M., Subramanian, S., Laird, E., and Kenny, R.A. (2020). Editorial: low population mortality from COVID-19 in countries south of latitude 35 degrees North supports vitamin D as a factor determining severity. Aliment. Pharmacol. Ther. 51, 1434–1437.

37. Tian, Y., Rong, L., Nian, W., and He, Y. (2020). Review article: gastrointestinal features in COVID-19 and the possibility of faecal transmission. Aliment Pharmacol Ther 51, 843–851.

38. Akalin, E., Azzi, Y., Bartash, R., Seethamraju, H., Parides, M., Hemmige, V., Ross, M., Forest, S., Goldstein, Y.D., Ajaimy, M., et al. (2020). Covid-19 and Kidney Transplantation. N. Engl. J. Med.

39. Alberici, F., Delbarba, E., Manenti, C., Econimo, L., Valerio, F., Pola, A., Maffei, C., Possenti, S., Zambetti, N., Moscato, M., et al. (2020). A single center observational study of the clinical characteristics and short-term outcome of 20 kidney transplant patients admitted for SARS-CoV2 pneumonia. Kidney Int.

40. Zhang, H., Chen, Y., Yuan, Q., Xia, Q.-X., Zeng, X.-P., Peng, J.-T., Liu, J., Xiao, X.-Y., Jiang, G.-S., Xiao, H.-Y., et al. (2020). Identification of Kidney Transplant Recipients with Coronavirus Disease 2019. Eur. Urol. 77, 742–747.

41. Lippi, G., Plebani, M., and Henry, B.M. (2020). Thrombocytopenia is associated with severe coronavirus disease 2019 (COVID-19) infections: A meta-analysis. Clin. Chim. Acta 506, 145–148.

42. Yang, X., Yang, Q., Wang, Y., Wu, Y., Xu, J., Yu, Y., and Shang, Y. (2020). Thrombocytopenia and its association with mortality in patients with COVID-19. J. Thromb. Haemost. 18, 1469–1472.

43. COVID-19 Host Genetics Initiative (2020). The COVID-19 Host Genetics Initiative, a global initiative to elucidate the role of host genetic factors in susceptibility and severity of the SARS-CoV-2 virus pandemic. Eur. J. Hum. Genet. 28, 715–718.

44. Ellinghaus, D., Degenhardt, F., Bujanda, L., Buti, M., Albillos, A., Invernizzi, P., Fernández, J., Prati, D., Baselli, G., Asselta, R., et al. (2020). Genomewide Association Study of Severe Covid-19 with Respiratory Failure. N. Engl. J. Med.

45. Nguyen, A., David, J.K., Maden, S.K., Wood, M.A., Weeder, B.R., Nellore, A., and Thompson, R.F. (2020). Human leukocyte antigen susceptibility map for SARS-CoV-2. J. Virol.

46. Blanco-Melo, D., Nilsson-Payant, B.E., Liu, W.-C., Uhl, S., Hoagland, D., Møller, R., Jordan, T.X., Oishi, K., Panis, M., Sachs, D., et al. (2020). Imbalanced Host Response to SARS-CoV-2 Drives Development of COVID-19. Cell 181, 1036-1045.e9.

47. Mancia, G., Rea, F., Ludergnani, M., Apolone, G., and Corrao, G. (2020). Renin-Angiotensin-Aldosterone System Blockers and the Risk of Covid-19. N. Engl. J. Med.

48. Reynolds, H.R., Adhikari, S., Pulgarin, C., Troxel, A.B., Iturrate, E., Johnson, S.B., Hausvater, A., Newman, J.D., Berger, J.S., Bangalore, S., et al. (2020). Renin-Angiotensin-Aldosterone System Inhibitors and Risk of Covid-19. N. Engl. J. Med.

49. Zhang, P., Zhu, L., Cai, J., Lei, F., Qin, J.-J., Xie, J., Liu, Y.-M., Zhao, Y.-C., Huang, X., Lin, L., et al. (2020). Association of Inpatient Use of Angiotensin Converting Enzyme Inhibitors and Angiotensin II Receptor Blockers with Mortality Among Patients With Hypertension Hospitalized With COVID-19. Circulation Research, CIRCRESAHA.120.317134.

50. D’Antiga, L. (2020). Coronaviruses and Immunosuppressed Patients: The Facts During the Third Epidemic. Liver Transpl.

51. Mehta, P., McAuley, D.F., Brown, M., Sanchez, E., Tattersall, R.S., Manson, J.J., and HLH Across Speciality Collaboration, UK (2020). COVID-19: consider cytokine storm syndromes and immunosuppression. Lancet 395, 1033–1034.

52. Monti, S., Balduzzi, S., Delvino, P., Bellis, E., Quadrelli, V.S., and Montecucco, C. (2020). Clinical course of COVID-19 in a series of patients with chronic arthritis treated with immunosuppressive targeted therapies. Ann Rheum Dis 79, 667–668.

53. Novi, G., Mikulska, M., Briano, F., Toscanini, F., Tazza, F., Uccelli, A., and Inglese, M. (2020). COVID-19 in a MS patient treated with ocrelizumab: does immunosuppression have a protective role? Mult Scler Relat Disord 42, 102120.

54. Ritchie, A.I., and Singanayagam, A. (2020). Immunosuppression for hyperinflammation in COVID-19: a double-edged sword? Lancet 395, 1111.

55. Shang, L., Zhao, J., Hu, Y., Du, R., and Cao, B. (2020). On the use of corticosteroids for 2019-nCoV pneumonia. Lancet 395, 683–684.

56. Zha, L., Li, S., Pan, L., Tefsen, B., Li, Y., French, N., Chen, L., Yang, G., and Villanueva, E.V. (2020). Corticosteroid treatment of patients with coronavirus disease 2019 (COVID-19). Med. J. Aust. 212, 416–420.

57. Tang, N., Bai, H., Chen, X., Gong, J., Li, D., and Sun, Z. (2020). Anticoagulant treatment is associated with decreased mortality in severe coronavirus disease 2019 patients with coagulopathy. J. Thromb. Haemost. 18, 1094–1099.

58. Thachil, J. (2020). The versatile heparin in COVID-19. J. Thromb. Haemost. 18, 1020–1022.

59. Thachil, J., Tang, N., Gando, S., Falanga, A., Cattaneo, M., Levi, M., Clark, C., and Iba, T. (2020). ISTH interim guidance on recognition and management of coagulopathy in COVID-19. J. Thromb. Haemost. 18, 1023–1026.

60. Little, P. (2020). Non-steroidal anti-inflammatory drugs and covid-19. BMJ 368, m1185.

61. Mehra, M.R., Ruschitzka, F., and Patel, A.N. (2020). Retraction—Hydroxychloroquine or chloroquine with or without a macrolide for treatment of COVID-19: a multinational registry analysis. The Lancet 0.

62. The Novel Coronavirus Pneumonia Emergency Response Epidemiology Team (2020). The Epidemiological Characteristics of an Outbreak of 2019 Novel Coronavirus Diseases (COVID-19) — China, 2020. CCDCW 2, 113–122.

63. Hirsch, J.S., Ng, J.H., Ross, D.W., Sharma, P., Shah, H.H., Barnett, R.L., Hazzan, A.D., Fishbane, S., Jhaveri, K.D., Northwell COVID-19 Research Consortium, et al. (2020). Acute kidney injury in patients hospitalized with COVID-19. Kidney Int.

64. Wang, D., Hu, B., Hu, C., Zhu, F., Liu, X., Zhang, J., Wang, B., Xiang, H., Cheng, Z., Xiong, Y., et al. (2020). Clinical Characteristics of 138 Hospitalized Patients With 2019 Novel Coronavirus-Infected Pneumonia in Wuhan, China. JAMA.

65. Huang, Y., and Zhao, N. (2020). Generalized anxiety disorder, depressive symptoms and sleep quality during COVID-19 outbreak in China: a web-based cross-sectional survey. Psychiatry Res 288, 112954.

66. Qiu, J., Shen, B., Zhao, M., Wang, Z., Xie, B., and Xu, Y. (2020). A nationwide survey of psychological distress among Chinese people in the COVID-19 epidemic: implications and policy recommendations. Gen Psychiatr 33.

67. Steinman, M.A., Perry, L., and Perissinotto, C.M. (2020). Meeting the Care Needs of Older Adults Isolated at Home During the COVID-19 Pandemic. JAMA Intern Med.

68. Lau, F.H., Majumder, R., Torabi, R., Saeg, F., Hoffman, R., Cirillo, J.D., and Greiffenstein, P. (2020). Vitamin D Insufficiency is Prevalent in Severe COVID-19. medRxiv, 2020.04.24.20075838.

69. CDC COVID-19 Response Team (2020). Severe Outcomes Among Patients with Coronavirus Disease 2019 (COVID-19) — United States, February 12–March 16, 2020. MMWR Morb Mortal Wkly Rep 69.

70. Garg, S., Kim, L., Whitaker, M., O’Halloran, A., Cummings, C., Holstein, R., Prill, M., Chai, S.J., Kirley, P.D., Alden, N.B., et al. (2020). Hospitalization Rates and Characteristics of Patients Hospitalized with Laboratory-Confirmed Coronavirus Disease 2019 - COVID-NET, 14 States, March 1-30, 2020. MMWR Morb. Mortal. Wkly. Rep. 69, 458–464.

71. Richardson, S., Hirsch, J.S., Narasimhan, M., Crawford, J.M., McGinn, T., Davidson, K.W., Barnaby, D.P., Becker, L.B., Chelico, J.D., Cohen, S.L., et al. (2020). Presenting Characteristics, Comorbidities, and Outcomes Among 5700 Patients Hospitalized With COVID-19 in the New York City Area. JAMA 323, 2052–2059.

72. Brandt, E.B., Beck, A.F., and Mersha, T.B. (2020). Air pollution, racial disparities, and COVID-19 mortality. J. Allergy Clin. Immunol.

73. Chowkwanyun, M., and Reed, A.L. (2020). Racial Health Disparities and Covid-19 - Caution and Context. N. Engl. J. Med.

74. Kuo, C.-L., Pilling, L.C., Atkins, J.L., Masoli, J.A.H., Delgado, J., Kuchel, G.A., and Melzer, D. (2020). APOE e4 genotype predicts severe COVID-19 in the UK Biobank community cohort. J. Gerontol. A Biol. Sci. Med. Sci.

75. Laurencin, C.T., and McClinton, A. (2020). The COVID-19 Pandemic: a Call to Action to Identify and Address Racial and Ethnic Disparities. J Racial Ethn Health Disparities 7, 398–402.

76. Ross, J., Diaz, C.M., and Starrels, J.L. (2020). The Disproportionate Burden of COVID-19 for Immigrants in the Bronx, New York. JAMA Intern Med.

77. Zhou, Z.-G., Xie, S.-M., Zhang, J., Zheng, F., Jiang, D.-X., Li, K.-Y., Zuo, Q., Yan, Y.-S., Liu, J.-Y., Xie, Y.-L., et al. (2020). Short-Term Moderate-Dose Corticosteroid Plus Immunoglobulin Effectively Reverses COVID-19 Patients Who Have Failed Low-Dose Therapy.

78. Fisher, R.A. (1922). On the interpretation of χ 2 from contingency tables, and the calculation of P. Journal of the Royal Statistical Society 85, 87–94.

79. McCullagh, P., and Nelder, J.A. (1989). Generalized Linear Models 2 edition. (Chapman and Hall/CRC).

80. Wu, P., Gifford, A., Meng, X., Li, X., Campbell, H., Varley, T., Zhao, J., Carroll, R., Bastarache, L., Denny, J.C., et al. (2019). Mapping ICD-10 and ICD-10-CM Codes to Phecodes: Workflow Development and Initial Evaluation. JMIR Med Inform 7, e14325.

81. Firth, D. (1993). Bias reduction of maximum likelihood estimates. Biometrika, 27–38.

82. Heinze, G. (2006). A comparative investigation of methods for logistic regression with separated or nearly separated data. Stat Med 25, 4216–4226.

83. Bonferroni, C.E., Bonferroni, C., and Bonferroni, C.E. (1936). Teoria statistica delle classi e calcolo delle probabilita’.

